# HPV T-cell epitope landscape: systematic mapping of distribution, conservation, and HLA promiscuity of known epitopes to inform immune-monitoring and vaccine design

**DOI:** 10.1101/2025.11.03.25339381

**Authors:** Syandrez Prima Putra, Selin Cankat, Leo Swadling

## Abstract

**Background:** Human papillomavirus (HPV) drives both malignant and benign tumors. Current prophylactic vaccines remain type-restricted, are not optimized for T-cell induction, and have not been efficacious therapeutically. T-cells can both block and control viral infections, supporting T-cell-based vaccines. We compile a comprehensive list of the currently known T-cell epitopes. We analyze their distribution and immunodominance and perform *in silico* analysis to further investigate conservation and HLA-promiscuity, to guide future epitope mapping, support immune monitoring, and to guide prophylactic and therapeutic pan-HPV vaccine design.

**Methods:** Functionally validated HPV T-cell epitopes were curated from the Immune Epitope Database (IEDB), filtered by length, HLA restriction, and immunogenicity. Epitopes were mapped across viral proteins, genotypes and HLAs. Additionally, we assessed conservation of each epitope across 454 distinct complete HPV genomes and predicted their HLA-promiscuity.

**Results:** 485 unique functionally validated HPV epitopes have been described (133 studies; 1,494 functional assays), originating from heterogeneous study contexts, including healthy donors, HPV-positive patients, and vaccination studies. Consistent with the study focus and viral biology, E6 and E7 proteins account for >60% of known HPV epitopes despite accounting for ∼10% of the viral proteome. L2 has so far yielded only one validated epitope despite being conserved. High-risk HPV types, especially HPV16 and HPV18, were the most studied (*p*<.001), and were enriched for CD8⁺ epitopes (OR 6.18; 95% CI 3.40–11.22; *p*<.001). Epitopes were restricted by 27 Class I and 21 Class II HLA alleles. *In silico* analysis indicated differences in epitope conservation between structural and non-structural proteins, and between high-risk and low-risk HPV types. Conserved, immunodominant, and HLA-promiscuous epitopes were highlighted from existing data.

**Conclusion:** This systematic analysis describes the dominance of E6/E7 in the reported HPV T-cell epitope landscape and quantitatively reveals major gaps in experimentally validated epitopes within conserved regions, particularly in L2. Addressing these gaps will be essential for a comprehensive assessment of cellular immunity and for rational next-generation HPV vaccine design.

## Introduction

Human papillomavirus (HPV) is a circular double-stranded DNA virus with an ∼8 kb genome and is responsible for approximately 5% of all human cancers, including cervical, vulvar, vaginal, penile, anal, and head and neck ^1^. HPV-related cervical cancer remains a leading cause of cancer-related mortality among women globally, especially in settings with limited access to vaccination and screening ^2^. HPV comprises more than 450 distinct genotypes when defined by <90% sequence identity in the L1 gene relative to other known types, of which 12 genotypes (HPV16, 18, 31, 33, 35, 39, 45, 51, 52, 56, 58, and 59) are classified as carcinogenic or high-risk types by the International Agency for Research on Cancer (IARC) ^3^. The development and global deployment of prophylactic bivalent, quadrivalent, and nonavalent L1-based virus-like particle vaccines has the potential to prevent 70-90% of HPV-related cancers associated with vaccine-covered genotypes ^4^. However, these vaccines are type-restricted, non-universal, and lack therapeutic efficacy against established infection or disease. In low- and middle-income countries, where HPV-associated cancers remain highly prevalent, vaccination coverage is limited by cost and infrastructure, and screening and treatment programs are often insufficient, resulting in a substantial residual disease burden even for vaccine-covered genotypes ^5,6^. Together, these limitations highlight the need for next-generation HPV vaccines that are cross-protective, affordable, and have both prophylactic and therapeutic applications.

One promising approach lies in leveraging T-cell immunity ^7–9^. Beyond neutralizing antibodies, T-cells play a crucial role in controlling viral infections by eliminating infected cells ^10^. Several lines of evidence point to a key role for T-cell immunity in controlling HPV infection and associated neoplasia ^11^. HPV-specific T-cells can be found in the blood and at higher numbers in the tumor site and draining lymph node in HPV-infected individuals ^9,12^. Those who progress to cervical intraepithelial neoplasia have a lower magnitude T-cell response than those who don’t ^12,13^. Expansion of HPV-specific T-cells has also been temporally correlated with regression of warts ^14^. Genetic association studies have repeatedly found associations between outcome and human leukocyte antigen (HLA), in particular for class II alleles ^15–17^ and HIV positivity and associated defects in cellular immunity are associated with higher rates of HPV acquisition, persistence and cancer progression ^18,19^.

We, and others, have recently shown that T-cells can protect against detectable viral infection even in the absence of antibodies ^20,21^, in particular memory T-cell responses targeting conserved viral regions, as observed in abortive SARS-CoV-2 infection ^22^. These observations establish a general immunological principle whereby T-cell immunity targeting conserved viral regions can in some circumstances be sufficient to mediate cross-variant and potentially infection-blocking protection. T-cells can recognize all viral proteins - including highly-conserved non-structural regions that are not accessible to antibodies, are inherently cross-reactive, and tend to be more long-lived than humoral immunity ^8,23,24^. This highlights the potential of T-cells to provide broad and more durable anti-viral protection. Applying this principle beyond SARS-CoV-2 provides a conceptual framework for HPV, whose viral proteome comprises early proteins (E1–E7), involved in replication and oncogenesis, and late proteins (L1, L2), forming the viral capsid ^25^. As with SARS-CoV-2, T-cell epitopes derived from conserved, potentially functionally constrained, regions of the HPV proteome may enable cross-genotype immunity by targeting regions that are less subject to immune escape ^22,26^. HLA molecules present viral peptides to T-cells, and their polymorphism determines population-level immunity as demonstrated in previous HPV studies ^27^. Comprehensive mapping of HPV T-cell epitopes across viral proteins, HPV types and HLA-restriction is therefore critical to identify the most promising candidates for broad-spectrum vaccine design.

To begin to address this gap, we established a systematic framework to analyze known HPV T-cell epitopes curated from published studies. The Immune Epitope Database (IEDB) provides the most comprehensive and standardized repository of experimentally validated T-cell epitopes (https://www.iedb.org) ^28^, enabling reproducible large-scale analyses across pathogens. Our focus was threefold: to chart known epitope distribution across proteins and genotypes, to evaluate conservation against a reference panel of complete genomes, and to assess HLA-promiscuity using *in silico* predictions. This approach enables an assessment of how extensively HPV T-cell epitopes have been explored to date, and where key gaps remain. It also acts as a resource for individuals designing reagents (peptide pools, MHC multimers) to investigate cellular immunity to HPV in natural history or vaccine studies. Ultimately, the framework aims to guide the prioritization of conserved and promiscuous epitopes as candidates for next-generation HPV vaccines, while also laying the groundwork for broader application to other oncogenic viruses.

## Methods

### Study design, data collection and initial curation strategy

To systematically characterize the currently reported and experimentally validated T-cell epitope landscape of HPV, we performed a retrospective meta-analysis of epitope data curated from the Immune Epitope Database (IEDB), complemented by *in silico* analyses (https://www.iedb.org; **Fig.1a**) ^28^. For the inclusion criteria, we searched the database using the following filters: (1) filter options = T-cell (excluding B-cell and MHC binding-only assays), (2) epitope = any, (3) epitope source, organism = Human papillomavirus (all types), (4) T-cell assay outcome = positive, (5) MHC restriction = any, (6) host = *Homo sapiens* (human), (7) disease = any, and (8) reference = any. We exported the results from the ‘Assays’ query as full data columns. To maintain consistency, we merged epitopes with identical peptide sequences across studies and counted each sequence once as a unique epitope for distribution analyses, while preserving all associated assay information and references. We then screened the peptides by length, retaining only 8–11 amino acids for Major Histocompatibility Complex (MHC) Class I restricted CD8⁺ T-cell epitopes and 12–23 amino acids for MHC Class II restricted CD4⁺ T-cell epitopes, and excluded those outside these ranges ^29^. We used this curated dataset as a core for analysis (485 epitopes, ‘proteome analysis’ epitope list). To minimize bias, screening and extraction were repeated three times.

**Fig. 1:**
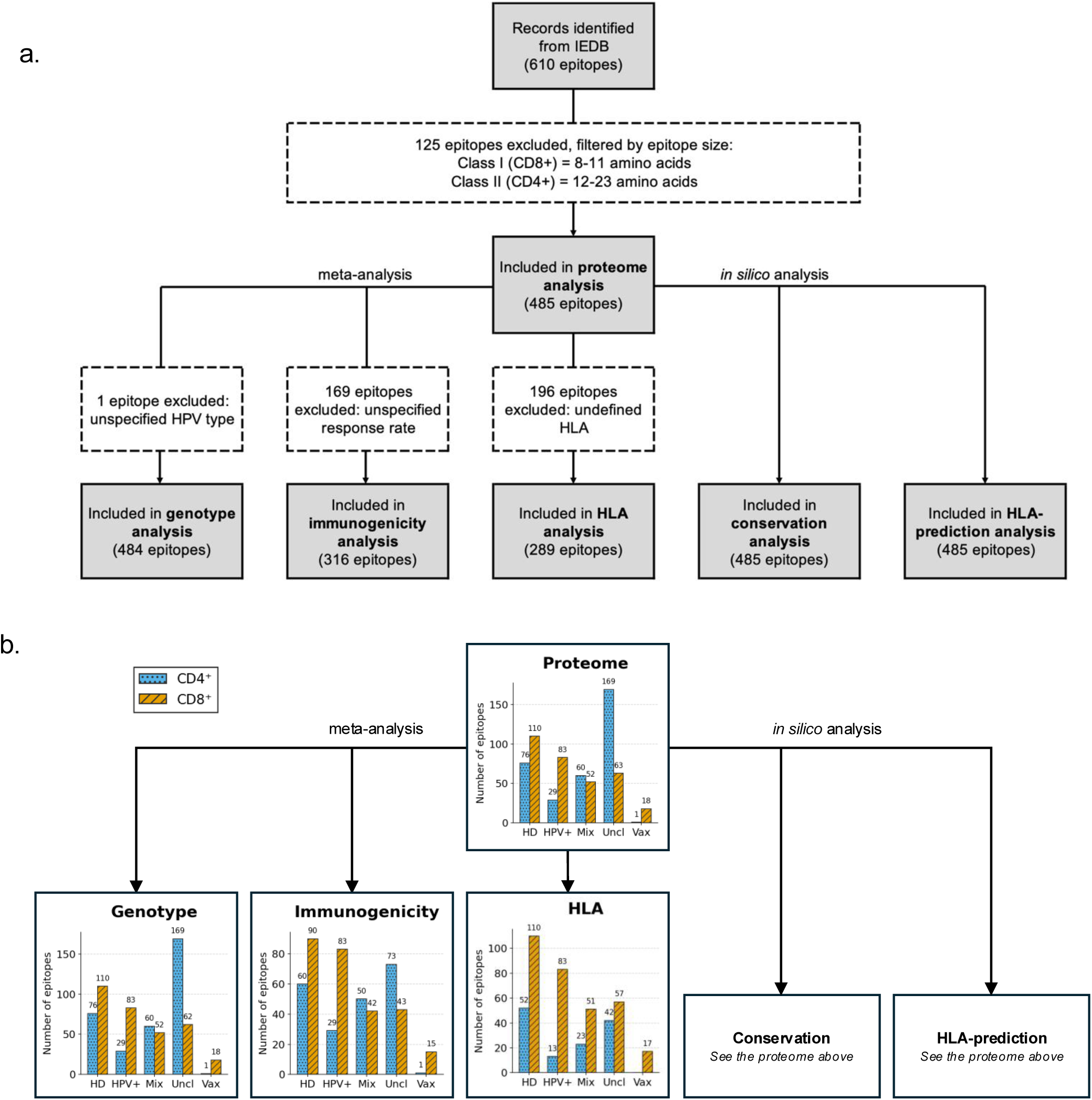
Data curation and study context of HPV T-cell epitopes. Workflow showing **(a)** identification and filtering of HPV T-cell epitopes from the Immune Epitope Database (IEDB) followed by meta- and in-silico analyses; and **(b)** distribution of curated epitopes across study contexts: healthy donors (HD), HPV-positive patients (HPV⁺), mixed/other patient groups (Mix), studies with incomplete annotation (Uncl), and vaccination studies (Vax). Data were obtained from IEDB (http://www.IEDB.org) based on queries on August 14th, 2025.

### Proteome analysis

We downloaded the HPV16 reference sequence FASTA file from NCBI GenBank (NC_001526.4) and extracted the protein annotations to construct a representative HPV proteome map, showing the relative positions and amino acid lengths of each protein. Using the IEDB core dataset, we then mapped all unique epitopes onto this proteome. Protein annotations were curated directly from the core dataset. Epitope density was calculated for each HPV protein (E6, E7, E1, E2, E4, E5, L2, and L1) as the number of epitopes per 100 amino acids to allow intuitive comparison across the relatively small HPV proteins. For cross-virus comparisons (e.g., SARS-CoV-2), values were normalized per 1,000 amino acids.

### Genotype analysis

From the ‘proteome analysis’ dataset, we filtered unique T-cell epitopes based on HPV genotype annotation and excluded epitopes with unspecified type from genotype analysis to give the ‘genotype analysis’ epitope list (n=484). HPV risk groups were classified according to the IARC, comprising 12 high-risk types (HPV16, 18, 31, 33, 35, 39, 45, 51, 52, 56, 58, and 59), with the remaining types designated as low risk. Epitopes with a single genotype annotation were classified as single-type, whereas those associated with two or more genotypes were classified as cross-type.

### Immunogenicity analysis

Immunogenicity here is defined as the reported response rate in IEDB, calculated as the percentage of positive responses across all subjects tested in functional assays compiled in IEDB. For each unique epitope, the total number of subjects tested and the number of positive responders from different studies were pooled, and the response rate was recalculated (range 0–1). Where the number of subjects tested and with positive responses were not specified in IEDB the epitopes were excluded, leaving 316 epitopes in the ‘immunogenicity’ epitope list. Subject-level response frequencies were compared between CD4⁺ and CD8⁺ T-cell epitopes and across HPV proteins. The proportional contribution of each HPV protein to the overall T-cell epitope pool was illustrated using stacked horizontal bar plots, showing the relative percentage and number (n) of epitopes per protein. Epitope response profiles were visualized using immunome browser–style plots (IEDB), where response rates were mapped against amino acid positions within each protein sequence.

### HLA analysis

Epitopes from the ‘proteome analysis’ dataset were filtered based on HLA (MHC class) annotation. All epitopes without defined HLA were excluded from the analysis, leaving 289 epitopes in the ‘HLA analysis’ epitope list. Epitope subsets were stratified into known MHC class I (CD8⁺) and MHC class II (CD4⁺) annotations according to IEDB data. Of these, 219 epitopes with allele-level HLA resolution were used for restriction analyses. The distribution of epitopes with known HLA annotation and allele-level restriction was presented in absolute numbers. To characterize HLA promiscuity, epitopes annotated with a single HLA allele were classified as mono-allelic, whereas those with more than one allele were classified as multi-allelic, or HLA-promiscuous epitope. Candidate epitopes for vaccine development were prioritized if they demonstrated a response rate >0.5 and were restricted by more than one HLA allele.

### Conservation analysis

To evaluate conservation, each unique T-cell epitope from the ‘proteome analysis’ dataset (n=485) was mapped *in silico* against 454 complete HPV genomes with known GenBank ID retrieved from the Papillomavirus Episteme (PaVE, https://pave.niaid.nih.gov/explore/reference_genomes/human_genomes; **Supp Table. S1**) ^30^. To assess conservation of epitopes across known diversity in viral sequence of HPV, we included all the reference genomes curated by PaVE. This dataset includes 228 genomes not classified by the HPV reference center but that have <90% conservation of L1 with all known HPV types and have been included in PaVE. 71 genomes pending approval, and the 155 reference genomes recognized by the HPV reference centre. We purposefully included reference and non-reference sequences from PaVe to include all known sequence variability. This allowed us to identify regions that have remained conserved as HPV has diversified, potentially uncovering functionally constrained regions to target with T-cell immunity. The tables provided can be sub-setted on only high-risk types if conservation just across the most clinically relevant sequences is needed.

Mapping was performed using six-frame translation to capture epitopes encoded on both forward and reverse strands. Conservation was defined as a 100% amino acid match. For downstream analyses, conservation was summarized at the HPV type level: epitope conservation was expressed as absolute counts (N) and percentages (%) of HPV types in which the epitope sequence was found identically across all 454 reference genomes. Conservation was further stratified by protein, peptide length, and T-cell type (CD4⁺ or CD8⁺). Statistical comparisons of conservation distributions were performed for proteins, T-cell subsets and risk groups. The top 30 HPV types with the highest number of mapped epitopes were identified, with annotation of their risk classification (high-risk vs low-risk). A proteome map was constructed to visualize the distribution of fully conserved (100%) epitopes across HPV proteins. The map was stratified by HPV risk group (high-risk vs low-risk) and further annotated according to T-cell subset (CD8⁺ or CD4⁺) and protein functional category (oncoprotein, replication, or structural).

### HLA prediction analysis

To evaluate HLA promiscuity, each unique epitope from the ‘proteome analysis’ dataset (n=485) was assessed for predicted HLA binding *in silico* using NetMHCpan (v4.1) ^31^ and NetMHCIIpan (v4.1) ^32^ following the default recommended thresholds. HLA supertypes group alleles with overlapping peptide-binding specificities, enabling population-level inference of T-cell epitope coverage using a limited set of representative molecules^33^. Binding affinity to MHC class I alleles (CD8⁺) was predicted across a panel of 20 representative HLA class I molecules, selected to represent the major HLA class I supertypes: HLA-C*02:02, HLA-A*02:01, HLA-C*04:01, HLA-A*03:01, HLA-A*01:01, HLA-A*11:01, HLA-C*06:02, HLA-B*08:01, HLA-A*26:01, HLA-C*03:03, HLA-C*03:04, HLA-C*01:02, HLA-B*15:01, HLA-A*24:02, HLA-B*58:01, HLA-B*39:01, HLA-B*07:02, HLA-B*40:01, HLA-A*33:03, and HLA-B*27:05 ^34^. These alleles were selected to balance broad population-level immunogenetic coverage with computational tractability. For MHC class II epitopes (CD4⁺), binding was predicted across the IEDB reference panel of HLA class II molecules (including HLA-DRB1*07:01, HLA-DQA1*01:02/DQB1*06:04, HLA-DQA1*02:01/DQB1*02:01, HLA-DRB1*09:01, HLA-DPA1*01:03/DPB1*02:01, HLA-DQA1*01:02/DQB1*05:01, HLA-DPA1*01:03/DPB1*04:01, HLA-DRB1*13:01, HLA-DRB1*0101, HLA-DQA1*01:03/DQB1*05:01, HLA-DRB1*03:01, HLA-DPA1*01:03/DPB1*04:02, HLA-DRB1*11:01, HLA-DRB1*15:01, HLA-DRB1*04:01, HLA-DQA1*04:01/DQB1*03:01, HLA-DQA1*05:01/DQB1*03:01, HLA-DQA1*05:05/DQB1*03:01, HLA-DRB1*04:03, and HLA-DQA1*01:02/DQB1*06:02) ^34^. Predicted binding affinity was expressed as percentile rank scores, with lower values indicating stronger binding. For MHC class I, epitopes were classified as strong binders (SB) when EL rank ≤0.5%, weak binders (WB) when 0.5–2%, and non-binders when >2%. For MHC class II, epitopes were classified as SB when rank ≤1%, WB when 1–5%, and non-binders when >5%. HLA binding predictions were used to assess relative HLA promiscuity and coverage rather than to infer functional immunogenicity. For each epitope, the number of predicted HLA binders (SB or WB) was calculated to quantify promiscuity. Epitope distributions were differentiated systematically for CD8⁺/MHC I and CD4⁺/MHC II subsets by: (1) non-binders versus binders, (2) mono- versus multi-allelic, (3) comparison of total HLA binders per epitope, (4) HPV proteins, and (5) the number of WB and SB in each HLA allele. Heatmaps were generated to depict the binding profiles of the top 25 promiscuous peptides within each HPV protein across the representative HLA allele panel.

### Statistical analysis and reproducibility

To ensure rigorous evaluation of the data, we applied statistical methods tailored to the type and distribution of each variable. Categorical outcomes were analyzed using Pearson’s chi-square test or Fisher’s exact test, and the strength of association was expressed as odds ratios (OR) with 95% confidence intervals (CI). For continuous variables that were not normally distributed, comparisons were performed with the Mann–Whitney U test or Kruskal–Wallis test. Correlations between variables were assessed using Spearman’s rank correlation (ρ). A p-value <0.05 was considered statistically significant. For immunogenicity, 95% CIs were estimated using the Wilson score method. All analyses were carried out in Python v3.12.11, employing established scientific libraries including pandas (v2.2.2), NumPy (v2.0.2), SciPy (v1.16.1), statsmodels (v0.14.5), matplotlib (v3.10.0), seaborn (v0.13.2), and Biopython (v1.85). Figures were produced in vector format to ensure publication quality.

## Results

### Defining a core dataset of HPV T-cell epitopes

To synthesise the known antigenic landscape of HPV and to identify knowledge gaps relevant to rational vaccine design and immune monitoring, we collated 610 peptide sequences reported across 133 human studies/1,494 different functional assays from the IEDB (**Fig.1a**). To ensure consistency in size, we excluded 125 peptides that did not meet peptide length criteria, resulting in a core dataset (‘proteome analysis’ epitope list) of 485 unique epitopes, consisting of 293 CD4⁺ and 192 CD8⁺ epitopes (**Supp Table. S2**). This curated dataset highlights both the diversity of epitope sources and the availability of high-quality, functionally validated sequences. Building on this curated dataset, we next explored six key aspects of HPV epitope biology: (1) proteome distribution, (2) genotype distribution, (3) immunogenicity, (4) HLA restriction, followed by *in silico* prediction of (5) epitope conservation and (6) HLA binding (**Fig.1a**). For genotype analysis, we retained 484 epitopes after excluding one with an unspecified HPV type. For immunogenicity, we analyzed 316 epitopes with available response data. For HLA analysis, we focused on 289 epitopes with clearly defined HLA. Finally, *in silico* prediction was performed on all 485 epitopes. Together, these refinements provide a robust foundation for dissecting epitope conservation, breadth of recognition, and their implications for T-cell–mediated protection.

The curated dataset comprises epitopes derived from diverse study contexts, including healthy donors, HPV-positive patients, other/mixed patients, vaccination studies, and incompletely annotated cohorts (**Fig. 1b**), reflecting the heterogeneous nature of the available HPV T-cell epitope literature. Epitopes described only in vaccine studies may in some cases reflect peptides that are not processed and presented during natural infection, however, these were a small subset of the known epitopes described here (19/485).

### Proteome-wide mapping highlights dominance of E6 and E7 epitopes

First, we explored the antigen distribution patterns of the 485 known unique functionally validated T-cell epitopes across the entire HPV proteome (**Fig.2a-d**). The proteome of HPV is composed of oncoproteins (E6, E7, E5), replication proteins (E1, E2, E4), and structural proteins (L1, L2)(**Fig.2a**). E8^E2-derived epitopes were not included in this analysis due to the absence of experimentally validated T-cell epitope data for this protein in the IEDB at the time of analysis. CD8⁺ epitopes were dominated by 9-mers, whilst CD4⁺ epitopes were largely 15-mers (**Supp Fig.1a**). The number of studies mapping epitopes in each viral protein differed remarkably: E6/E7 were the most assayed proteins, with L2 being represented in just two studies (**Fig.2b**). E6 contributed the most epitopes for both CD8⁺ and CD4⁺ T-cells (70 and 90, respectively), followed by E7 (60 and 48) and L1 (16 and 77). In contrast, only a single epitope has been described in L2 (**Fig.2c**), despite L2 being identified as a promising candidate antigen for vaccines due to its conserved regions ^35^. Many CD4⁺ T-cell epitopes have been identified in the main target of currently licensed vaccines, L1, likely due to an interest in CD4⁺ help for these antibody-based vaccines.

**Fig. 2:**
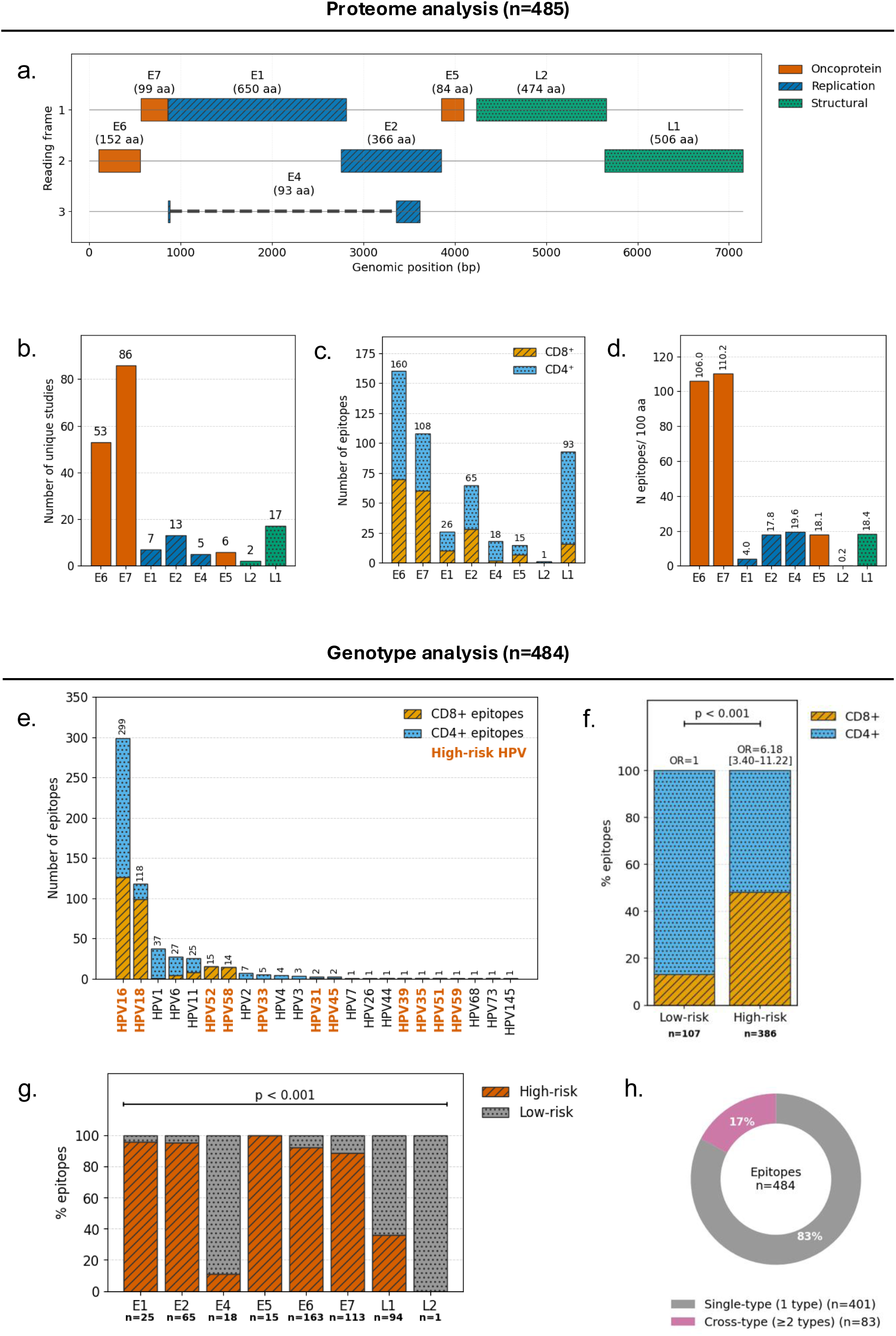
Proteome-wide mapping and genotype-level distribution of HPV T-cell epitopes. Proteome analysis of 485 HPV T-cell epitopes with defined peptide length and protein annotations **(a-d)**, showing: **(a)** genomic map of HPV proteins by reading frame (HPV-16; Ref.Seq: NC_001526.4), colored by functional class (oncoprotein E5, E6, E7=vermillion, replication E1, E2, E4=navy-hatched, structural L1, L2=green-dotted) with amino acid lengths indicated and dashed lines connecting spliced segments; **(b)** number of studies reporting epitopes per protein; **(c)** number of epitopes per protein based on T-cell subsets; and **(d)** epitope density, showing the number of known epitopes per 100 amino acids of each protein. Genotype analysis of 484 HPV T-cell epitopes with defined genotype annotations **(e-h)**, illustrating: **(e)** the number of epitopes per HPV type stratified by T-cell subset (CD8⁺ =yellow-hatched, CD4⁺=light blue-dotted), IARC-defined high-risk types are highlighted in bold orange; **(f)** proportion of epitopes restricted to high-risk versus low-risk groups, with p-value and odds ratio (OR, 95% CI) shown (reference=low-risk); **(g)** distribution of epitopes across HPV proteins (E1–L2) stratified by risk group (high-risk=vermilion-hatched, low-risk=gray-dotted) with global p-value indicated; and **(h)** proportion of epitopes restricted to a single HPV type (gray) versus those conserved across multiple types (cross-type epitopes, pink).

To account for the difference in length of viral proteins epitope density was calculated as the number of epitopes per 100 amino acids length. The density of known epitopes varied widely by protein and no significant association was observed between protein length and epitope density across the HPV proteome (ρ=−0.52, *p*=0.183) (**Fig.2d**, **Supp Fig.1b**). Instead, T-cell epitope representation was dominated by the small oncoproteins E6 and E7, which emerged as clear outliers with disproportionately high epitope density, while other similarly sized proteins, such as E4 and E5, showed substantially lower epitope density. To contextualise this pattern, we applied the epitope density analysis to SARS-CoV-2 T-cell epitopes, which have been extensively and systematically mapped^36^. Epitope density for SARS-CoV-2 proteins fell between ∼100-300 epitopes per 1,000aa over a wide range of protein lengths, without pronounced outliers like E6/E7. Proteins L2 and E1 have a lower number of known epitopes than would be expected for their size (**Supp Fig.1b**), and they are understudied (**Fig.2b**), highlighting them as candidate proteins for future epitope mapping. Together, these findings may indicate that current HPV epitope datasets are strongly shaped by research focus on E6/E7, but that there is also likely an additional contribution from underlying viral biology, such as high and prolonged protein expression for these oncoproteins.

### High-risk types of HPV are overrepresented

To explore how genotype and clinical risk categories influence distribution among reported T-cell epitopes, we compared 484 unique HPV epitopes with known genotype information (**Fig.2e-h**). Most epitopes were identified in studies of high-risk HPV types. Significantly more epitopes were identified in studies of HPV16 and HPV18 than low-risk types (**Fig.2e**). Known epitopes selected from high-risk types were approximately six times as likely to be CD8⁺ as CD4⁺ (OR 6.18; 95% CI 3.40–11.22; *p*<.001) (**Fig.2f**). When stratified by protein, high-risk types were represented with increased epitope representation in E5, E2, E1, E6, and E7 but reduced epitope representation in L1, L2, and E4, compared to low-risk (*p*<.001) (**Fig.2g**). This bias is expected as there is much interest in the therapeutic potential of cytotoxic CD8⁺s targeting the oncoproteins of the high-risk genotypes. While most epitopes were reported for a single genotype, approximately 17% were found to be present among multiple types of HPV (**Fig.2h**). Very few studies assessed the conservation of epitopes or the cross-reactivity of T-cell responses across genotypes and so this is a significant underestimation of the potential for pan-genotype epitopes. Below we compile a reference sequence dataset and perform *in silico* analysis to better assess epitope conservation. These data overall reflect a high scientific emphasis upon high-risk types of HPV, particularly CD8⁺-restricted epitopes, but reveal shared epitopes as promising candidates to generate broad-spectrum vaccines.

### Immunodominance of E6 and E7 across and within HPV protein landscape

To better understand the immunogenic regions within HPV proteins, rather than looking at the epitope density, we consider the response rates to known epitopes (the proportion of exposed individuals with the correct HLA type who make a response to the peptide). We analyzed 316 reported epitope T-cell responses, examined the frequency of positive responses among tested subjects according to T-cell types (CD4⁺ and CD8⁺) and proteins, and then mapped them to the viral proteome (**Fig.3a-b; Supp Fig.1c-d**).

**Fig. 3:**
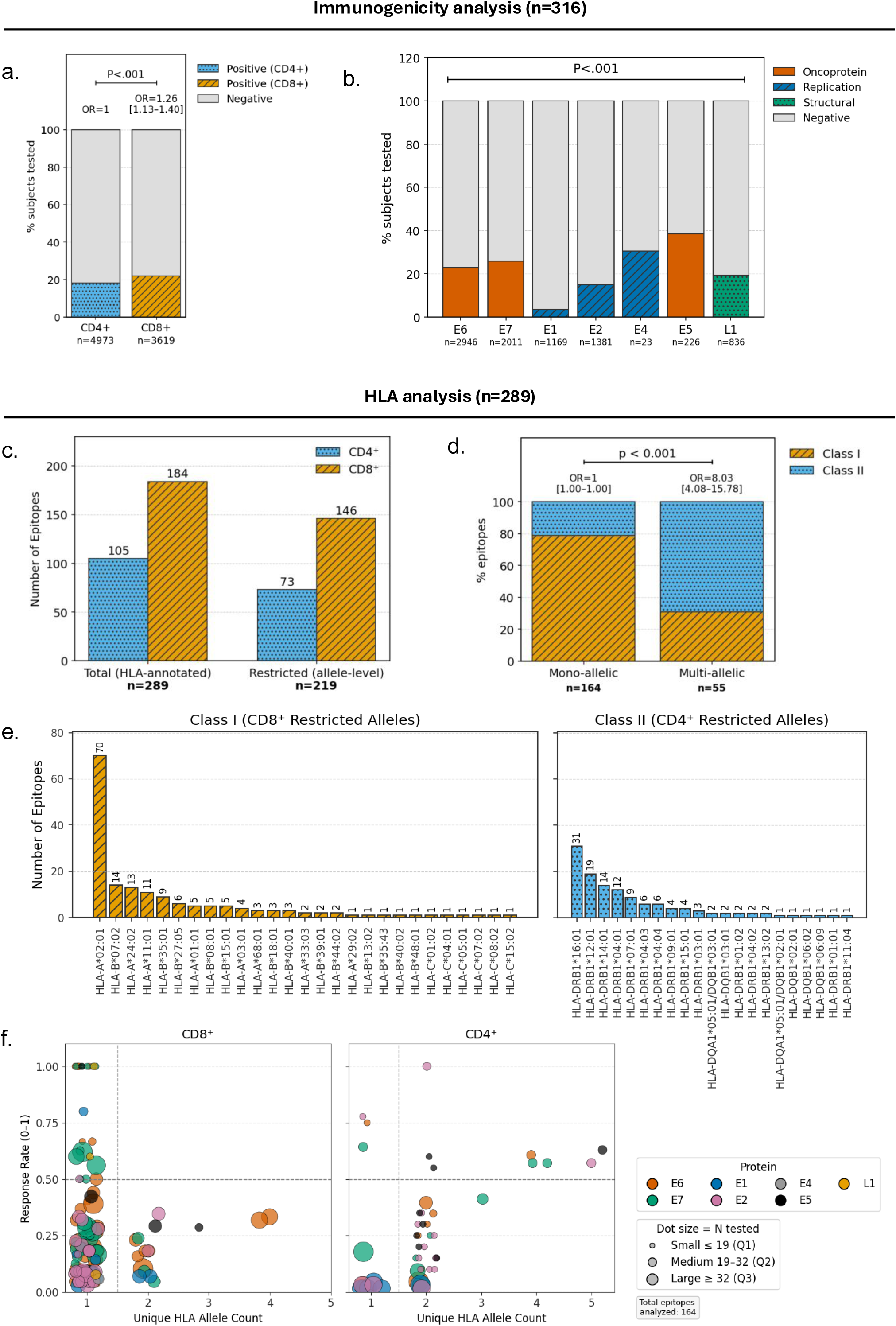
HLA restriction landscape and immunogenicity patterns of HPV T-cell epitopes. Immunogenicity analysis of 316 HPV T-cell epitopes with defined response rates **(a-b),** showing: **(a)** proportion of subjects tested with CD4⁺ (light blue-dotted) and CD8⁺ (yellow-hatched) epitopes showing positive (colored) versus negative (gray) immune responses, with p value and OR [95% CI] indicated; **(b)** proportion of subjects tested with epitopes from oncoprotein E5-E7 (vermillion), replication E1-E4 (navy blue-hatched) and structural L1-L2 (green-dotted) protein epitopes showing positive (colored) versus negative (gray) immune responses, with global p value and OR [95% CI] indicated. Analysis of 289 epitopes with defined HLA-annotation and 219 allele-level-restricted **(c-f)**, illustrating: **(c)** number of CD4⁺ (light blue-dotted) and CD8⁺ (yellow-hatched) epitopes across all HLA-annotated (total) and allele-level restricted datasets, with total n indicated below each group; **(d)** proportion of class I (yellow-hatched) and Class II (light blue-dotted) epitopes classified as mono-allelic or multi-allelic (HLA-promiscuous) binders, with OR [95% CI] and p value indicated; **(e)** number of class I (CD8⁺, yellow-hatched) and class II (CD4⁺, light blue-dotted) restricted epitopes mapped to individual HLA alleles; **(f)** correlation between response rate and HLA promiscuity among CD8⁺ and CD4⁺ epitopes. Each dot represents a unique epitope, colored by HPV protein and scaled by the number of subjects tested (dot size, quartiles shown in legend).

CD8⁺ responses were more frequently detected than CD4⁺ responses (OR=1.26, 95%CI=1.13-1.40, *p*<.001) (**Fig.3a**). Oncoproteins E5, E6 and E7 elicited higher response rates on average for known epitopes, whereas early proteins like E1 and E2 were less immunogenic (*p*<.001) (**Fig.3b**). To investigate whether regions with higher immunogenicity could be identified, response rates were plotted across the viral proteins (**Supp Fig.1d).** For several proteins there was not sufficient known epitopes to clearly see response rate patterns (E1, E2, L1, L2), however, immunogenic regions in E6, E7 and E5 could be identified (**Supp Fig.1c)**.

### Asymmetric HLA restriction highlights uneven allele coverage

To assess the potential breadth of T-cell cross-reactivity in genetically heterogeneous populations, we examined the HLA restriction profiles of HPV epitopes with available allele-level data. Of the 289 HLA-annotated epitopes identified, 219 had allele-level resolution and were therefore included in the restriction analysis (**Fig.3c-f; Supp Fig.2**). A larger number of epitopes with HLA annotation and restricting alleles have been identified (**Fig.3c**; **Supp Table. S3**). CD4⁺ epitopes were eight times more likely to be restricted by multiple alleles (promiscuous) than CD8⁺ epitopes (OR=8.03; 95% CI=4.08-15.78; *p*<0.001) (**Fig.3d**). Overall, HLA restriction profiles were unevenly distributed, with a small number of alleles accounting for most epitopes described for MHC class I and MHC class II (**Fig.3e**). A total of 27 Class I and 21 Class II HLA alleles restricted known HPV epitopes, with HLA-A*02:01 (70 epitopes) and HLA-DRB1*16:01 (31 epitopes). The number of epitopes identified per HLA allele did not differ significantly between Class I and Class II molecules (p=0.536) (**Supp Fig.2a**). The number of HLAs restricting epitopes within a protein were significantly correlated with the number of epitopes described for that protein, with E6 and E7 also being outliers for breadth of HLA-restriction, as they were for epitope density (**Supp Fig2.b-c**).

It may be desirable to include immunodominant, with high response rates, and HLA promiscuous peptides for global vaccines. We next explored the relationship between HLA allele restriction and response rates (**Fig.3f**). The most broadly restricted CD4⁺ epitope binds 5 different HLA alleles, compared to a maximum of 4 for CD8⁺ epitopes. Several promiscuous and immunodominant CD4 epitopes were identified. The most promiscuous epitopes were LLSVSTYTSLILLVL (E5_33-47_, response rate of 0.63) and GLYYVHEGIRTYFVQ (E2_156-170,_ response rate of 0.57) for CD4⁺. For CD8⁺, KLPDLCTEL (E6_13-21_, response rate of 0.33) and EYRHYCYSL (E6_75–83_, response rate of 0.32) were the most promiscuous but they had relatively low recorded response rates. These findings suggest that even though some HPV epitopes can bind to several HLA alleles, the current dataset remains shaped by uneven allele coverage and differential epitope mapping across viral proteins, rather than providing a comprehensive representation of HLA-restricted T-cell recognition.

### *In silico* prediction suggests limited cross-genotype conservation

It is crucial to understand how conserved the HPV epitopes are across genotypes to facilitate identification of candidates with broad-spectrum T-cell immunity potential. To pursue this further, we performed an *in silico* screen against a database of 454 HPV reference genomes (see methods) for the 485 functionally validated known epitopes (**Fig.4**; **Supp Table. S4**). CD8⁺ peptides generally covered more HPV types than CD4⁺ peptides. The maximum coverage of HPV T-cell epitopes was 34 (7.5%) genotypes for CD8⁺ epitope LQFIFQLCK (9-mer) and 21 (4.6%) for CD4⁺ epitope FNKPYWLQRAQGHNN (15-mer) (**Fig.4a**). Although overall conservation was low, structural proteins showed slightly higher conservation (median 0.44%) than non-structural proteins (0.22%, *p*<0.001) (**Fig.4b**). There was no statistical distinction evident between overall CD4⁺ versus CD8⁺ epitope conservation (**Supp Fig.3a**). At the genotype level, epitope matches were contributed mostly by HPV16 (n=288, 59.4%), followed by HPV6 (n=42, 8.7%), then by HPV1 (n=37, 8.1%) and HPV18 (n=36, 7.4%) (**Fig.4c**). High-risk and low-risk HPV epitopes showed similar median conservation (0.22% vs 0.22%), although the distributions differed (*p*<0.001) (**Fig.4d**). Conserved epitopes were unevenly distributed across the HPV proteome, with dense clusters observed in E6/E7 and the structural protein L1, and replication proteins harbored relatively few epitopes (**Fig.4e**). Overall, these results indicate that most experimentally reported HPV epitopes are highly genotype-specific and exhibit low cross-genotype conservation.

**Fig. 4:**
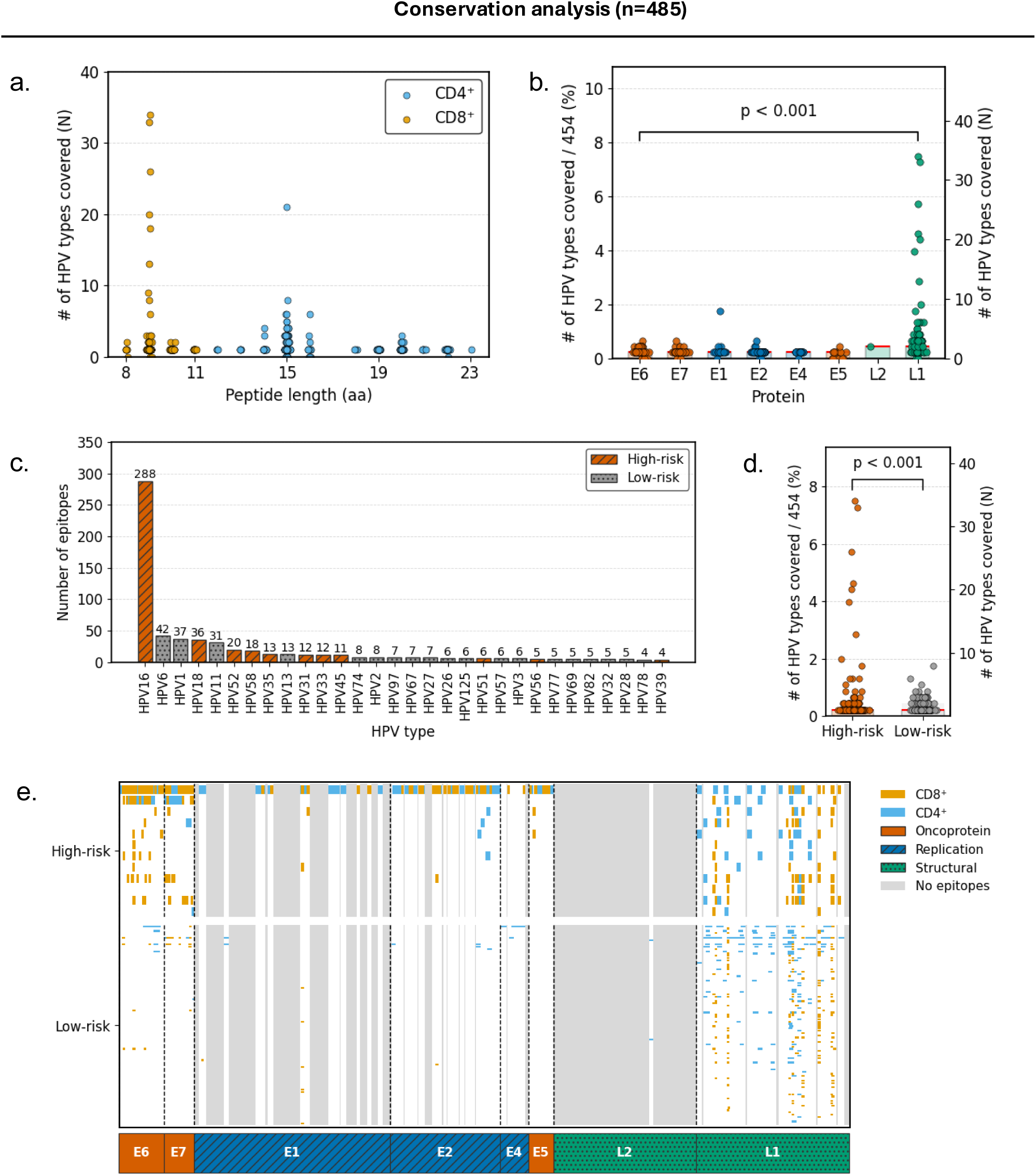
Epitope conservation across HPV genomes following *in silico* analysis. Conservation analysis of 485 unique T-cell epitopes towards 454 reference genomes, showing: **(a)** distribution of CD8⁺ (yellow) and CD4⁺ (light blue) epitopes by peptide length and number of HPV types covered; **(b)** cross-type coverage of unique epitopes across HPV proteins; **(c)** number of unique T-cell epitopes across HPV genotypes, grouped by high-risk (orange) and low-risk (gray) types; **(d)** proportion and number of HPV types covered by each epitope, compared between high-risk (orange) and low-risk (gray) groups; and **(e)** epitope distribution across the HPV proteome, showing the localization of CD8⁺ (yellow) and CD4⁺ (light blue) T-cell epitopes in high-risk and low-risk types, grouped by functional protein classes (oncoprotein, replication, and structural).

### Supertype-based HLA prediction framework enables concordance assessment and epitope prioritisation across the HPV dataset

To systematically assess HLA-binding breadth beyond single-allele annotations which are common in IEDB, we predicted binding of known HPV epitopes to a curated panel of alleles, selected as globally common alleles that represent the major HLA supertypes, using NetMHCpan and NetMHCIIpan (**Fig.5**; **Supp Fig.4**; **Supp Table. S5**). 219 epitopes had reported HLA restriction, with 119 CD8⁺ and 26 CD4⁺ epitopes eligible for direct concordance analysis based on allele representation within our supertype panel (**Supp Fig.4a–b**). Across the full dataset, CD8⁺ epitopes were more frequently predicted to bind at least one allele within the panel compared to CD4⁺ epitopes, both among reported (*p*=0.0019) and non-reported HLA subsets (*p*<.001) (**Fig.5a**). Within the allele-represented subset, concordance between predicted and reported alleles was significantly higher for CD8⁺ than CD4⁺ epitopes (*p*=0.0096) (**Fig.5b–c**), with allele-level analysis indicating that some mismatches reflected alternative predicted binders within related supertypes (**Supp Fig.4c–f**).

**Fig. 5:**
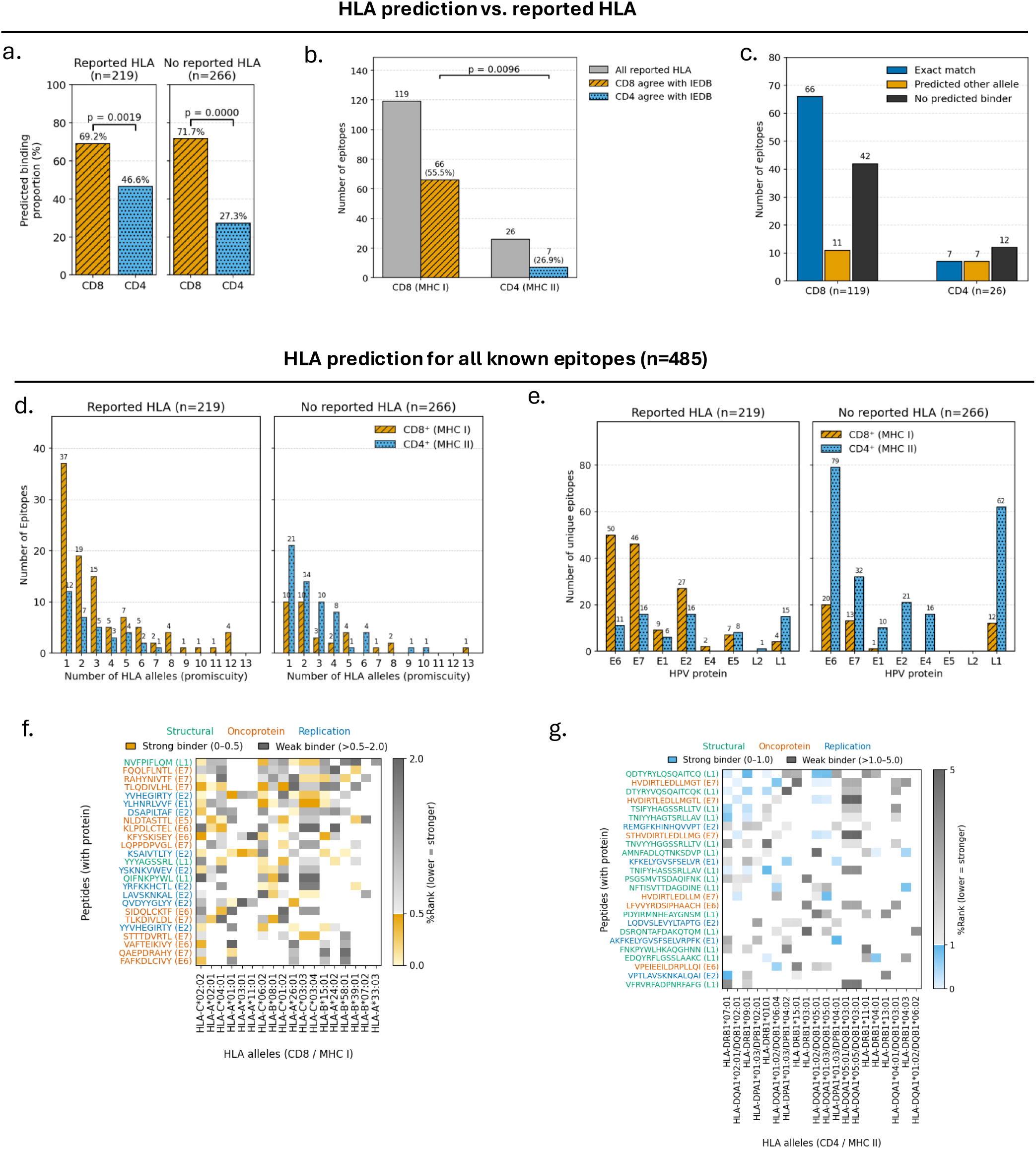
Supertype-based HLA binding prediction and epitope promiscuity analysis across HPV T-cell epitopes. *In silico* prediction of HLA binding for 485 validated HPV T-cell epitopes was performed using a curated 20-allele supertype panel (NetMHCpan and NetMHCIIpan). **(a)** Proportion of predicted binders among CD8⁺ and CD4⁺ epitopes, stratified by presence or absence of reported HLA restriction. **(b–c)** Concordance analysis between predicted binding and IEDB-reported HLA restriction within the allele-represented subset, including exact matches, alternative predicted binders, and non-predicted binders. **(d)** Distribution of predicted HLA promiscuity, defined as the number of binding alleles per epitope within the 20-allele panel, shown separately for epitopes with and without reported HLA restriction. **(e)** Protein-level distribution of predicted CD8⁺ and CD4⁺ epitopes across HPV proteins, stratified by reported versus non-reported HLA subsets. **(f–g)** Heatmaps of the top 25 most promiscuous CD8⁺ (Class I) and CD4⁺ (Class II) epitopes, illustrating predicted binding strength across the supertype panel. Strong binders are shown in color shades (yellow for CD8^+^ and light blue for CD4^+^) and weak binders in gray shades.

Predicted HLA promiscuity was next analysed separately for epitopes with reported (n=219) and without reported HLA restriction (n=266) (**Fig.5d–e**). CD8⁺ epitopes more frequently demonstrated multi-allelic binding, with some peptides predicted to bind up to 12 alleles in the reported HLA subset and up to 13 alleles in the non-reported subset, whereas CD4⁺ epitopes were generally restricted to fewer alleles (**Fig.5d**). Protein-level stratification showed that predicted CD8⁺ epitopes were most frequently observed in E6 and E7 across both subsets. In contrast, CD4⁺ epitopes were more evenly distributed across proteins in the reported HLA subset but showed relative enrichment in E6 and L1 in the non-reported HLA subset (**Fig.5e**). We visualised the top 25 most promiscuous CD8⁺ and CD4⁺ epitopes using heatmaps to illustrate predicted multi-allelic binding patterns across the supertype panel (**Fig.5f–g**). These potentially highly promiscuous peptides were derived from multiple HPV proteins, although their distribution remained uneven across the proteome. At the allele level, HLA-C*02:02 and HLA-A*02:01 accounted for the highest number of predicted class I binders, while HLA-DRB1*07:01 and HLA-DQA1*02:01/DQB1*02:01 accounted for the highest number of class II binders (**Supp Fig.4e–f**). Together, these findings support the utility of the supertype framework for identifying potentially broadly presented HPV T-cell epitopes while highlighting class-specific differences in predictive performance.

## Discussion

HPV is a major health priority, causing large numbers of anogenital and oropharyngeal cancers globally despite effective prophylactic vaccines. Current vaccines, though remarkably effective against vaccine-included types, offer limited cross-type coverage without therapeutic effect against pre-existing infections, leaving a critical void for disease attenuation ^37^. There is increasing evidence that T-cell immunity can act early to abort viral infections before they are established ^21,38^ as well as mediating viral clearance during established infections, making T-cell epitopes central targets for next-generation prophylactic vaccines as well as immunotherapeutics. Prophylactic approaches should benefit by moving beyond a narrow focus on neutralizing antibodies, to additionally exploiting the durability and cross-reactive potential of T-cell immunity. There are also many settings in which accurate quantification and assessment of the T-cell response to HPV may aid in our understanding of pathogenesis and correlates of protection. Systematic epitope mapping throughout the entire HPV proteome is required to aid the design of reagents for the measurement of HPV-specific T-cell immunity and to inform design of vaccine immunogens. In this context, our analyses provide a framework for prioritising conserved and HLA-promiscuous HPV regions that remain experimentally unexplored, illustrating how systematic *in silico* approaches can extend beyond the current IEDB knowledge base.

T-cells are not all equally effective at anti-viral control. Specificity (at the level of TCR, epitope, protein and virus) and immunodominance (both in terms of magnitude of the response and proportion of individuals recognizing an epitope) influences how effective a T-cells is and when a T-cell can act within an infection. An in-depth understanding of the locations of epitopes and response rates is essential. Our study brings together all currently available data on anti-HPV T-cell specificity and immunodominance, highlighting key knowledge gaps, and using *in silico* tools to start to fill these gaps.

To the best of our knowledge, this study offers the first systematic proteome-wide analysis of known HPV T-cell epitopes that has been curated and thoroughly annotated by protein, genotype, and HLA restriction. Extending prior reports of HPV epitopes ^39–42^. Comparable efforts in alternate viral systems, i.e., influenza virus and SARS-CoV-2 ^43,44^, have demonstrated the value of systematic epitope compilation and underscore the importance of such an enterprise for HPV.

Given the heterogeneous study contexts represented in the IEDB, including healthy donors, HPV-infected patients, and vaccinated individuals, the observed epitope landscape reflects a combination of underlying viral biology and prior experimental focus, rather than a comprehensive measure of intrinsic immunogenicity across the HPV proteome. This interplay itself is informative, revealing substantial gaps in epitope mapping across the HPV proteome, particularly for conserved structural and replication-associated proteins (e.g. L1 and E1).

HPV oncogenes E6 and E7 were highly represented among the epitope datasets, with peak epitope density reported despite their small size. This enrichment is consistent with long-standing evidence that E6 and E7 are frequent targets of host T-cell targets, uniformly preserved throughout tumor cells in expression profiles, and major causes of HPV-containing cancers ^45–49^. Notably, independent immunopeptidomics and expression-level studies have similarly reported predominant MHC presentation of peptides derived from E6/E7, with only occasional detection of other early proteins such as E1 ^9^, supporting a biological basis for their prominence beyond epitope discovery efforts alone. Epitopes within E6/E7 were skewed toward CD8⁺ T-cell responses and high-risk HPV genotypes, a pattern that has informed the development of some prospective therapeutic vaccine candidates targeting these proteins ^50–52^. At the same time, this distribution also reflects historical research prioritisation of E6/E7, particularly in the context of cancer immunotherapy. In contrast, proteins such as L2, have been investigated in fewer studies, translating to fewer known epitopes, despite accumulating evidence supporting their cross-type protective potential ^53–55^. Our previous work, which revealed minimal variation in HPV16 L2 sequences from Indonesian cervical cancer samples, supported the notion that L2 contains conserved regions worthy of further study ^35^. We also highlighted enrichment of CD4⁺ epitopes within L1, consistent with the extensive study of this protein in the context of licensed prophylactic antibody-based vaccines. Together, these observations indicate that the current HPV T-cell epitope landscape reflects a combination of biological antigen presentation patterns and historical study focus, rather than providing a comprehensive measure of intrinsic immunogenicity across the HPV proteome.

It is evident that T-cell epitope coverage in literature is comprised mostly of high-risk HPV types with a special focus on HPV16, consistent with its established ranking among the most common genotypes associated with cancers ^25,56^. The importance of identifying and exploiting CD8⁺ epitopes in oncogenes of high-risk genotypes is supported by previous work showing diminished CD8⁺ function is accompanied by impaired viral clearance and higher risk for cancers ^57–59^. Protein targets that vary among high- compared to low-risk types also indicate a focus in previous research on early proteins central to viral replication as well as malignant conversion ^60^. Of particular interest is the identification of epitopes that span multiple types of HPV to signify their importance as cross-protective antigens for a vaccine, which has already been implemented among several viruses, such as influenza ^26^ and SARS-CoV-2 ^61^.

Importantly, analysis of HLA restriction revealed that HPV epitopes had broad coverage of a large repertoire of Class I and Class II molecules, however, they were concentrated disproportionately within a restricted subset of alleles, most notably HLA-A*02:01 and HLA-DR4. Such distribution mirrors an extensive experimental focus on more common alleles as has been described for other viruses ^34,62^. The closed groove of MHC class I enforces stricter binding specificity, while the open-ended groove of MHC class II permits longer and more variable peptides (Rock et al., 2016). This is reflected in a preponderance towards mono-allelic restriction amongst CD8⁺ epitopes suggests narrower coverage of individual CD8⁺ epitopes than CD4⁺ epitopes, with a concern for population coverage within highly disparate genetic backgrounds ^34^.

Our *in silico* analyses demonstrate that only a small percentage of known HPV T-cell epitopes exhibit broad conservation across types, with the majority being highly genotype-specific. In accordance with evidence that capsid-derived regions contain conserved epitopes that can induce cross-protection across diverse HPV types, structural proteins like L1 and L2 emerged as relatively richer sources of cross-type candidates ^53,63^. Although conserved fragments within E6/E7 have been reported and are being investigated as multigenotype therapeutic targets, epitopes from early proteins tended to be type-restricted ^64,65^.

Parallel HLA binding prediction showed that while most epitopes were predicted to bind a limited number of alleles, whereas a small subset demonstrated broad multi-allelic binding. These epitopes are especially useful because HLA promiscuity extends the range of individuals who can present and target the epitope with T-cell immunity ^66,67^. These findings underscore the dual challenge of limited conservation and restricted allele coverage for most HPV epitopes. Although no pan-genotype epitopes were identified here, we have highlighted a small number of epitopes with high response rates (immunodominant) and wide HLA restriction (promiscuous) for further investigation. Overall, these data show proof-of-concept for identifying HLA-promiscuous epitopes but call for systematic validation of HLA promiscuity and epitope conservation.

There are various limitations to this study. First, although one aim of this analysis was to identify knowledge gaps in epitope mapping, these inherent biases limit our ability to accurately determine immunodominance at the protein level and to fully characterize HLA promiscuity and cross-genotype conservation. But through highlighting these gaps we hope to inform future efforts. Second, although informative, *in silico* predictions of HLA binding and conservation cannot completely replace experimental confirmation of T-cell responses in various human populations. We only considered an epitope to be pan-genotypic if sequence identity was 100%, however, T-cells have flexibility in recognition and cross-reactivity can be retained to epitope variants. Cross-reactivity and potential for cross-protection is therefore underestimated when using this strict cutoff and functional cross-recognition of variants should be performed in vitro. Third, the analyses were limited to 454 genomes. These sequences have been curated by PaVE to include International HPV reference center recognized viral types and additional putative types with <90% sequence identity to these types, in order to reflect the known papillomavirus diversity. Using single references for each type may not accurately represent intra-type variation and the true diversity of HPV. Despite the controlled curation of IEDB and further filtering on epitopes that have been observed using functional T-cell assays, there is heterogeneity in the way epitopes have been identified and a reliance on reported annotations. Lastly, the evidence that is currently available may limit generalizability across the entire HPV spectrum by overemphasizing high-risk genotypes, especially HPV16. These drawbacks illustrate how crucial it is to combine methodical *in silico* techniques with more extensive experimental research to improve epitope selection and direct the creation of vaccines that are applicable to both populations and other types.

## Conclusion

In conclusion, this study provides the first proteome-wide map of HPV T-cell epitopes, integrating information on distribution, conservation, HLA restriction, and immunogenicity. Our findings reveal the focus on E6 and E7, the underrepresentation of structural proteins such as L2, and the limited conservation and HLA coverage that constrain most epitopes. At the same time, the identification of conserved and HLA-promiscuous sequences highlights promising candidates for cross-type and broadly protective vaccines. By bridging prophylactic and therapeutic perspectives, these results offer a rational framework to expand HPV vaccine design beyond neutralizing antibodies, emphasizing the potential of T-cell–based immunity to address both prevention of new infections and clearance of established disease.

## Data Availability

Data and Code Availability
All data analysed during this study are included in this published article and its Supplementary Information. Correspondence and requests for materials should be addressed to L.S. Custom scripts are available on request.

---

**Supp Fig. 1:**
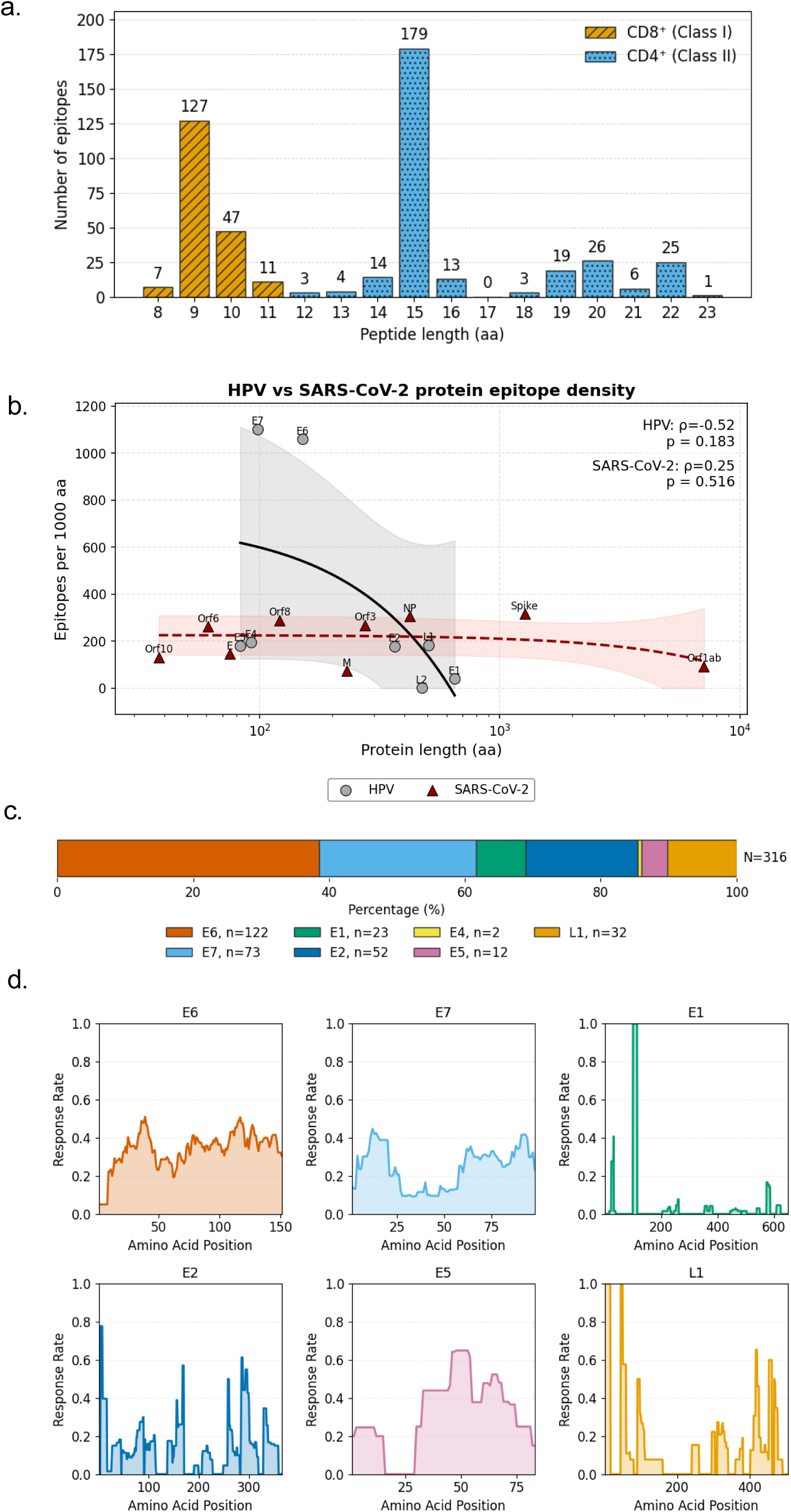
HPV T-cell epitopes length and immunodominance. Proteome analysis of 485 HPV T-cell epitopes with defined peptide length and protein annotations, showing: **(a)** number of experimentally validated CD8⁺ (Class I) and CD4⁺ (Class II) T-cell epitopes per HPV protein; and **(b)** Relationship between protein length and T-cell epitope density (epitopes per 1,000 amino acids) across HPV and SARS-CoV-2 proteins. Solid lines indicate linear regression with shaded areas representing 95% confidence intervals. Immunogenicity analysis of 316 HPV T-cell epitopes with defined response rates, illustrating: **(c)** distribution of epitopes with available response rate data, showing proportional representation of HPV proteins; and **(d)** immunogenicity profiles (response rate) across amino acid positions of individual HPV proteins, highlighting distinct regions of epitope clustering.

**Supp Fig. 2:**
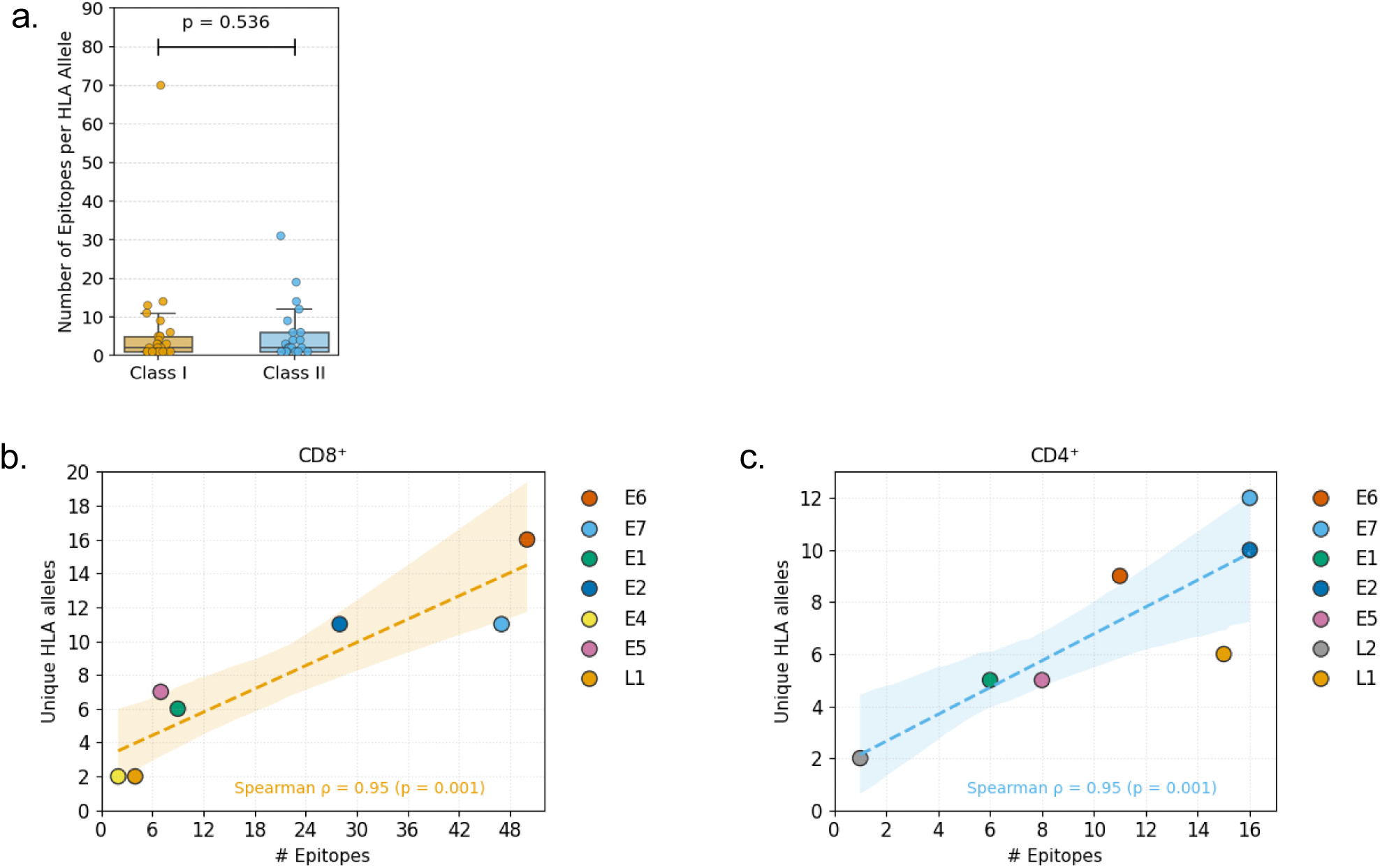
Relationship between HLA restriction breadth and epitope count. Analysis of 219 HLA allele-restricted illustrating: **(a)** comparison of the number of epitopes per HLA allele between Class I (CD8⁺) and Class II (CD4⁺) restrictions; and **(b–c)** correlations between the number of unique HLA alleles and total epitopes per protein for CD8⁺ (b, orange) and CD4⁺ (c, light blue) T-cell epitopes, with Spearman’s ρ and p-values indicated.

**Supp Fig. 3:**
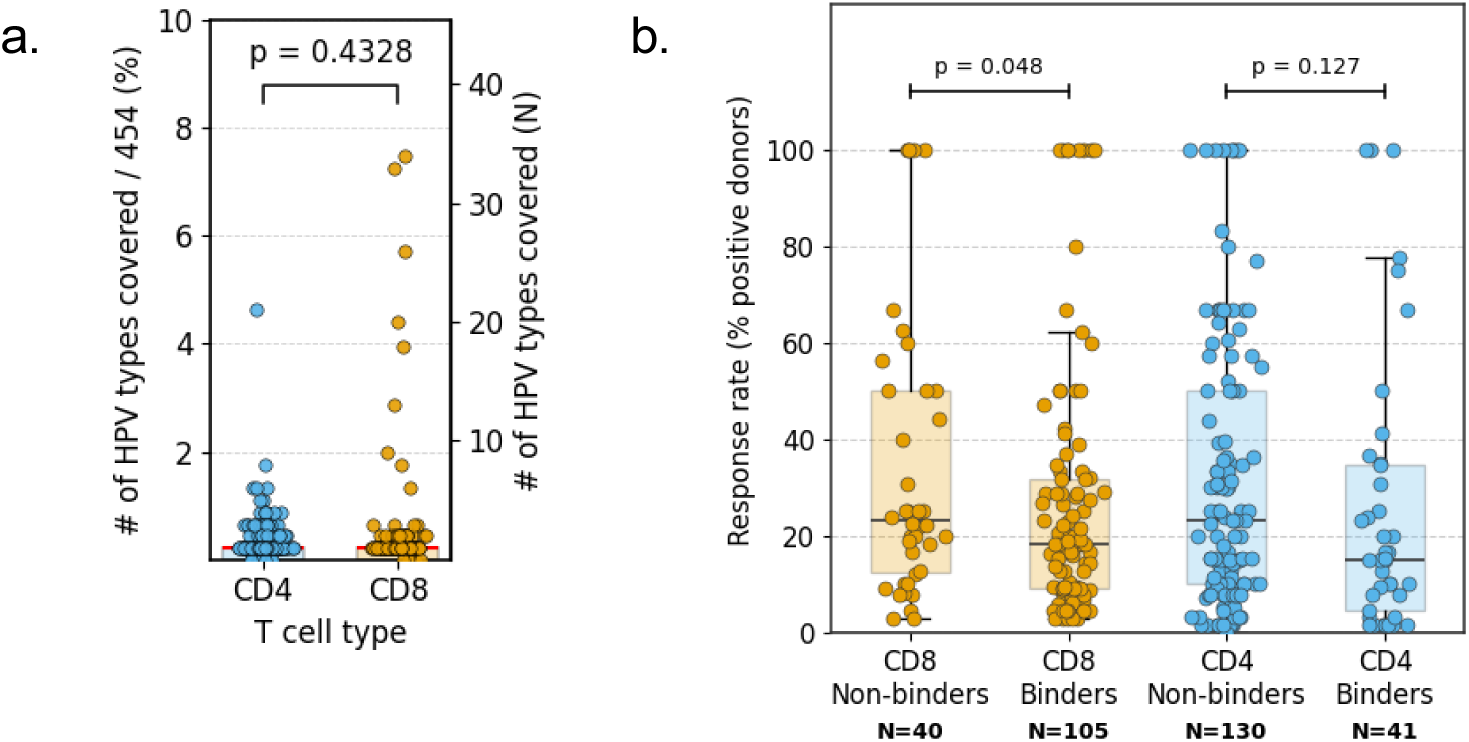
*In silico* analysis of HPV T-cell epitope type coverage and donor response rate by T-cell subset and binding status. Analysis of 485 unique T-cell epitopes mapped to 454 reference genomes showing **(a)** comparison of HPV types coverage between CD4⁺ and CD8⁺ T-cell epitopes, where each dot represents a unique epitope, and horizontal lines show medians with interquartile ranges. Analysis of 316 epitopes across 20 HLA-restricted alleles for both MHC Class I (CD8⁺) and Class II (CD4⁺) T-cells, showing **(b)** response rate (% positive donors) comparison between binders and non-binders.

**Supp Fig. 4:**
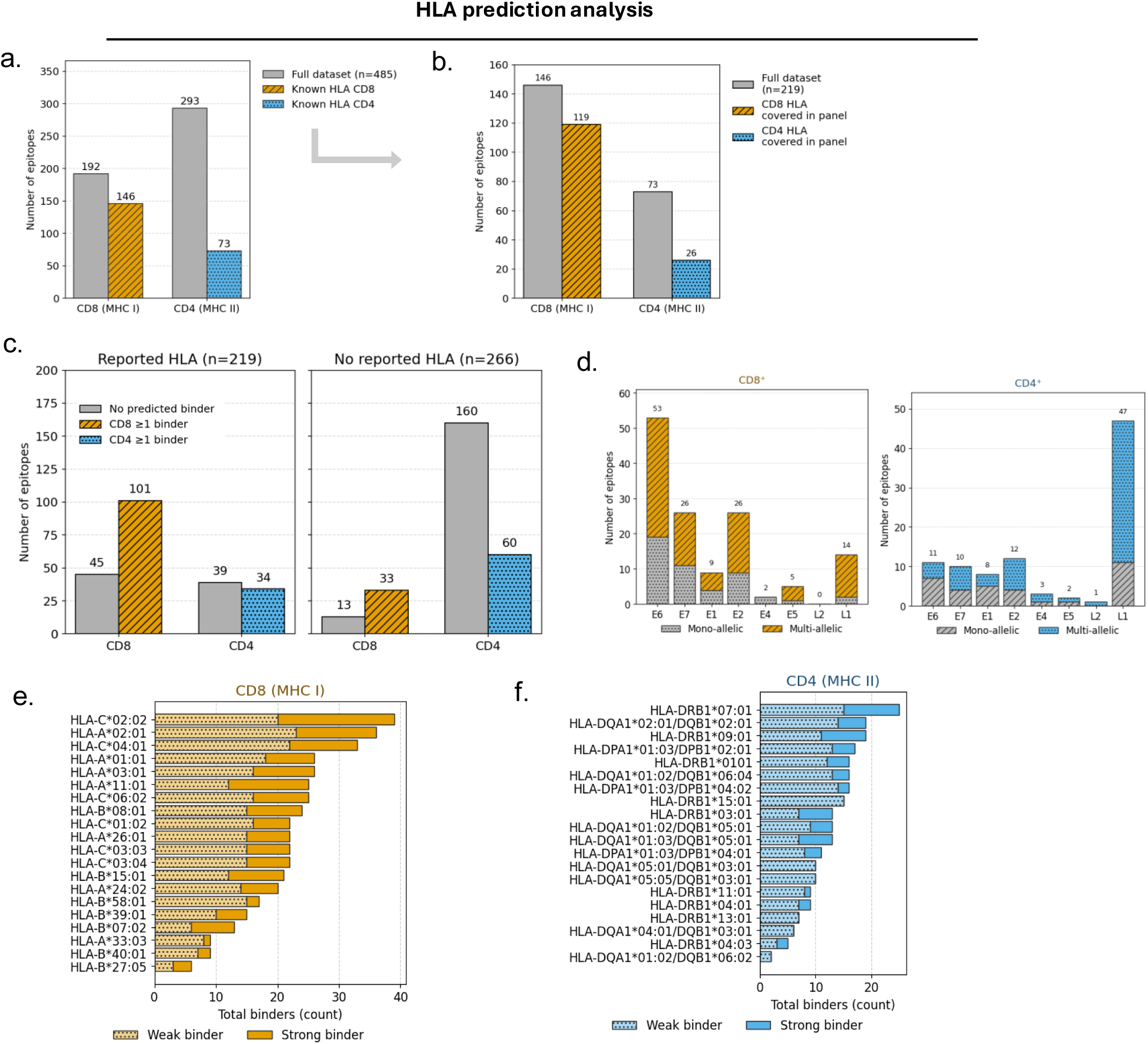
Framework and allele-level characteristics of supertype-based HLA prediction analysis. **(a)** Overview of the HPV T-cell epitope dataset (n=485), showing the distribution of CD8⁺ (MHC I) and CD4⁺ (MHC II) epitopes and the subset with reported HLA restriction. **(b)** Representation of reported HLA-restricted epitopes within the curated 20-allele supertype panel, indicating the number of CD8⁺ and CD4⁺ epitopes eligible for concordance analysis. **(c)** Predicted binding outcomes among epitopes with and without reported HLA restriction, stratified by CD8⁺ and CD4⁺ T-cell class, showing predicted binders versus non-binders within the panel. **(d)** Distribution of mono-allelic and multi-allelic predicted binders across HPV proteins for CD8⁺ and CD4⁺ epitopes. **(e–f)** Allele-level distribution of predicted binding counts across the 20-allele supertype panel for Class I (CD8⁺) and Class II (CD4⁺), respectively, indicating total predicted binders per allele separated into strong and weak binding categories.

**Supplementary Table S1:** 454 HPV genomes with accession number retrieved from PaVE database.

**Supplementary Table S2:** Meta-analysis of experimentally validated HPV T-cell epitopes with immunogenicity, HLA restriction, and assay information.

**Supplementary Table S3:** List of experimentally validated HPV T-cell epitopes restricted by specific HLA.

**Supplementary Table S4:** *In silico* analysis of conservation and HLA-binding profiles of HPV T-cell epitopes.

**Supplementary Table S5:** *In silico* analysis of HPV epitope binders across HLA alleles.

## Acknowledgements

We would like to thank Professor Hans Stauss and all members of the Swadling laboratory (UCL) for helpful discussions.

## Ethical approval

This study did not involve living subjects and thus was exempted from institutional ethical review board approval.

## Author Contributions

SPP and LS designed the study. SPP and SC collected and analysed data. SPP and LS drafted manuscripts. LS validated the methodology and verified the data. All authors read and approved the final manuscript.

## References

1. Baba, S.K., Alblooshi, S.S.E., Yaqoob, R., Behl, S., Al Saleem, M., Rakha, E.A., et al. (2025). Human papilloma virus (HPV) mediated cancers: an insightful update. J. Transl. Med. 23, 483. 10.1186/s12967-025-06470-x.

2. Sung, H., Ferlay, J., Siegel, R.L., Laversanne, M., Soerjomataram, I., Jemal, A., et al. (2021). Global Cancer Statistics 2020: GLOBOCAN estimates of incidence and mortality worldwide for 36 cancers in 185 countries. CA Cancer J. Clin. 71, 209–249. 10.3322/caac.21660.

3. Wei, F., Georges, D., Man, I., Baussano, I., and Clifford, G.M. (2024). Causal attribution of human papillomavirus genotypes to invasive cervical cancer worldwide: a systematic analysis of the global literature. Lancet 404, 435–444. 10.1016/S0140-6736(24)01097-3.

4. Wang, R., Pan, W., Jin, L., Huang, W., Li, Y., Wu, D., et al. (2020). Human papillomavirus vaccine against cervical cancer: opportunity and challenge. Cancer Lett. 471, 88–102. 10.1016/j.canlet.2019.11.039.

5. Boldeanu, L., Assani, M.-Z., Boldeanu, M.V., Siloși, I., Manolea, M.-M., Văduva, C.-C., et al. (2025). Cervical Cancer in the era of HPV: translating molecular mechanisms into preventive public health action. Int. J. Mol. Sci. 26, 8463. 10.3390/ijms26178463.

6. Tsu, V.D., LaMontagne, D.S., Atuhebwe, P., Bloem, P.N., and Ndiaye, C. (2021). National implementation of HPV vaccination programs in low-resource countries: lessons, challenges, and future prospects. Prev. Med. 144, 106335. 10.1016/j.ypmed.2020.106335.

7. Dull, P.M., Achilles, S.L., Ahmed, R., Barnabas, R.V., Campos, N.G., Chirgwin, K., et al. (2024). Meeting report: considerations for trial design and endpoints in licensing therapeutic HPV16/18 vaccines to prevent cervical cancer. Vaccine 42, 126100. 10.1016/j.vaccine.2024.07.001.

8. Eberhardt, C.S., Kissick, H.T., Patel, M.R., Cardenas, M.A., Prokhnevska, N., Obeng, R.C., et al. (2021). Functional HPV-specific PD-1+ stem-like CD8 T cells in head and neck cancer. Nature 597, 279–284. 10.1038/s41586-021-03862-z.

9. Peng, X., Woodhouse, I., Hancock, G., Parker, R., Marx, K., Müller, J., et al. (2023). Novel canonical and non-canonical viral antigens extend current targets for immunotherapy of HPV-driven cervical cancer. iScience 26, 106101. 10.1016/j.isci.2023.106101.

10. Huber, S.R., van Beek, J., de Jonge, J., Luytjes, W., and van Baarle, D. (2014). T cell responses to viral infections: opportunities for peptide vaccination. Front. Immunol. 5, 1–12. 10.3389/fimmu.2014.00001.

11. Van Bockel, D., and Kelleher, A. (2023). The crossroads: divergent roles of virus-specific CD4+ T lymphocytes in determining the outcome for human papillomavirus infection. Immunol. Cell Biol. 101, 525–534. 10.1111/imcb.12650.

12. Hilders, C.G.J.M., Ras, L., van Eendenburg, J.D.H., Nooyen, Y., and Fleuren, G.J. (1994). Isolation and characterization of tumor-infiltrating lymphocytes from cervical carcinoma. Int. J. Cancer 57, 805–813. 10.1002/ijc.2910570608.

13. Evans, E.M., Man, S., Evans, A.S., and Borysiewicz, L. (1997). Infiltration of cervical cancer tissue with human papillomavirus-specific cytotoxic T-lymphocytes. Cancer Res. 57, 2943–2950.

14. Coleman, N., Birley, H.D.L., Renton, A.M., Hanna, N.F., Ryait, B.K., Byrne, M., et al. (1994). Immunological events in regressing genital warts. Am. J. Clin. Pathol. 102, 768–774. 10.1093/ajcp/102.6.768.

15. Apple, R.J., Erlich, H.A., Klitz, W., Manos, M.M., Becker, T.M., and Wheeler, C.M. (1994). HLA DR–DQ associations with cervical carcinoma show papillomavirus-type specificity. Nat. Genet. 6, 157–162. 10.1038/ng0294-157.

16. Bontkes, H.J., Van Duin, M., De Gruijl, T.D., Duggan-Keen, M.F., Walboomers, J.M.M., Stukart, M.J., et al. (1998). HPV 16 infection and progression of cervical intra-epithelial neoplasia: analysis of HLA polymorphism and HPV 16 E6 sequence variants. Int. J. Cancer 78, 166–171. 10.1002/(SICI)1097-0215(19981005)78:2<166::AID-IJC8>3.0.CO;2-X.

17. Bowden, S.J., Bodinier, B., Kalliala, I., Zuber, V., Vuckovic, D., Doulgeraki, T., et al. (2021). Genetic variation in cervical preinvasive and invasive disease: a genome-wide association study. Lancet Oncol. 22, 548–557. 10.1016/S1470-2045(21)00028-0.

18. Clifford, G.M., Tully, S., and Franceschi, S. (2017). Carcinogenicity of human papillomavirus (HPV) types in HIV-positive women: a meta-analysis from HPV infection to cervical cancer. Clin. Infect. Dis. 64, 1228–1235. 10.1093/cid/cix135.

19. Stelzle, D., Tanaka, L.F., Lee, K.K., Ibrahim Khalil, A., Baussano, I., Shah, A.S.V., et al. (2021). Estimates of the global burden of cervical cancer associated with HIV. Lancet Glob. Health 9, e161–e169. 10.1016/S2214-109X(20)30459-9.

20. Fumagalli, V., Ravà, M., Marotta, D., Di Lucia, P., Bono, E.B., Giustini, L., et al. (2024). Antibody-independent protection against heterologous SARS-CoV-2 challenge conferred by prior infection or vaccination. Nat. Immunol. 25, 633–643. 10.1038/s41590-024-01787-z.

21. Kalimuddin, S., Tham, C.Y.L., Chan, Y.F.Z., Hang, S.K., Kunasegaran, K., Chia, A., et al. (2025). Vaccine-induced T cell responses control Orthoflavivirus challenge infection without neutralizing antibodies in humans. Nat. Microbiol. 10, 374–387. 10.1038/s41564-024-01903-7.

22. Swadling, L., Diniz, M.O., Schmidt, N.M., Amin, O.E., Chandran, A., Shaw, E., et al. (2022). Pre-existing polymerase-specific T cells expand in abortive seronegative SARS-CoV-2. Nature 601, 110–117. 10.1038/s41586-021-04186-8.

23. Dan, J.M., Mateus, J., Kato, Y., Hastie, K.M., Yu, E.D., Faliti, C.E., et al. (2021). Immunological memory to SARS-CoV-2 assessed for up to 8 months after infection. Science 371, eabf4063. 10.1126/science.abf4063.

24. Manisty, C., Treibel, T.A., Jensen, M., Semper, A., Joy, G., Gupta, R.K., et al. (2021). Time series analysis and mechanistic modelling of heterogeneity and sero-reversion in antibody responses to mild SARS-CoV-2 infection. EBioMedicine 65, 103259. 10.1016/j.ebiom.2021.103259.

25. Nelson, C.W., and Mirabello, L. (2023). Human papillomavirus genomics: understanding carcinogenicity. Tumour Virus Res. 15, 200258. 10.1016/j.tvr.2023.200258.

26. Eickhoff, C.S., Terry, F.E., Peng, L., Meza, K.A., Sakala, I.G., Van Aartsen, D., et al. (2019). Highly conserved influenza T cell epitopes induce broadly protective immunity. Vaccine 37, 5371–5381. 10.1016/j.vaccine.2019.07.033.

27. Nakagawa, M., Kim, K.H., Gillam, T.M., and Moscicki, A.-B. (2007). HLA class I binding promiscuity of the CD8 T-cell epitopes of human papillomavirus type 16 E6 protein. J. Virol. 81, 1412–1423. 10.1128/JVI.01768-06.

28. Vita, R., Blazeska, N., Marrama, D., Shackelford, D., Zalman, L., Foos, G., et al. (2025). The Immune Epitope Database (IEDB): 2024 update. Nucleic Acids Res. 53, D436–D443. 10.1093/nar/gkae1092.

29. Grifoni, A., Mahajan, S., Sidney, J., Martini, S., Scheuermann, R.H., Peters, B., et al. (2019). A survey of known immune epitopes in the enteroviruses strains associated with acute flaccid myelitis. Hum. Immunol. 80, 923–929. 10.1016/j.humimm.2019.08.004.

30. Dommer, J., Van Doorslaer, K., Afrasiabi, C., Browne, K., Ezeji, S., Kim, L., et al. (2025). PaVE 2.0: behind the scenes of the papillomavirus episteme. J. Mol. Biol. 437, 168925. 10.1016/j.jmb.2024.168925.

31. Reynisson, B., Alvarez, B., Paul, S., Peters, B., and Nielsen, M. (2020). NetMHCpan-4.1 and NetMHCIIpan-4.0: improved predictions of MHC antigen presentation by concurrent motif deconvolution and integration of MS MHC eluted ligand data. Nucleic Acids Res. 48, W449–W454. 10.1093/nar/gkaa379.

32. Reynisson, B., Barra, C., Kaabinejadian, S., Hildebrand, W.H., Peters, B., and Nielsen, M. (2020). Improved prediction of MHC II antigen presentation through integration and motif deconvolution of mass spectrometry MHC eluted ligand data. J. Proteome Res. 19, 2304–2315. 10.1021/acs.jproteome.9b00874.

33. Sidney, J., Peters, B., Frahm, N., Brander, C., and Sette, A. (2008). HLA class I supertypes: a revised and updated classification. BMC Immunol. 9, 1. 10.1186/1471-2172-9-1.

34. Sanchez-Mazas, A., Nunes, J.M., Di, D., Dominguez, E.A., Gerbault, P., Faye, N.K., et al. (2024). The most frequent HLA alleles around the world: a fundamental synopsis. Best Pract. Res. Clin. Haematol. 37, 101559. 10.1016/j.beha.2024.101559.

35. Putra, S., Putra, A., Astuti, I., and Asmara, W. (2022). Human papillomavirus type 16 L2 gene sequence variation analysis in Indonesian cervical cancer specimens. Asian Pac. J. Cancer Prev. 23, 2009–2016. 10.31557/APJCP.2022.23.6.2009.

36. Grifoni, A., Sidney, J., Vita, R., Peters, B., Crotty, S., Weiskopf, D., et al. (2021). SARS-CoV-2 human T cell epitopes: adaptive immune response against COVID-19. Cell Host Microbe 29, 1076–1092. 10.1016/j.chom.2021.05.010.

37. Mo, Y., Wang, Y., Zhang, L., Li, H., and Liu, J. (2022). Prophylactic and therapeutic HPV vaccines: current scenario and perspectives. Front. Cell. Infect. Microbiol. 12, 909223. 10.3389/fcimb.2022.909223.

38. Swadling, L., and Maini, M.K. (2023). Can T cells abort SARS-CoV-2 and other viral infections? Int. J. Mol. Sci. 24, 4371. 10.3390/ijms24054371.

39. Gallagher, K.M.E., and Man, S. (2007). Identification of HLA-DR1- and HLA-DR15-restricted human papillomavirus type 16 (HPV16) and HPV18 E6 epitopes recognized by CD4+ T cells from healthy young women. J. Gen. Virol. 88, 1470–1478. 10.1099/vir.0.82576-0.

40. McInnis, C., Wylie, B., Grulich, A.E., Pyeon, D., and Frazer, I.H. (2023). Identification of HPV16 E1- and E2-specific T cells in the oropharyngeal cancer tumor microenvironment. J. Immunother. Cancer 11, e006721. 10.1136/jitc-2023-006721.

41. Mercier-Letondal, P., Martel, C., Côté, C., Charbonneau, B., Roy, J., and Perreault, C. (2018). Isolation and characterization of an HLA-DRB1*04-restricted HPV16-E7 T cell receptor for cancer immunotherapy. Hum. Gene Ther. 29, 1202–1212. 10.1089/hum.2018.041.

42. Speetjens, F.M., Kenter, G.G., van Poelgeest, M.I.E., Welters, M.J.P., and Melief, C.J.M. (2022). Intradermal vaccination of HPV-16 E6 synthetic peptides conjugated to an optimized Toll-like receptor 2 ligand shows safety and potent T cell immunogenicity in patients with HPV-16 positive (pre-)malignant lesions. J. Immunother. Cancer 10, e005016. 10.1136/jitc-2022-005016.

43. Chaisawangwong, W., Mueangtam, P., Mongkolsapaya, J., and Screaton, G.R. (2022). Cross-reactivity of SARS-CoV-2- and influenza A-specific T cells in individuals exposed to SARS-CoV-2. JCI Insight 7, e158308. 10.1172/jci.insight.158308.

44. Gaevert, J.A., Luque Duque, D., Lythe, G., Molina-París, C., and Thomas, P.G. (2021). Quantifying T cell cross-reactivity: influenza and coronaviruses. Viruses 13, 1786. 10.3390/v13091786.

45. Inturi, R., and Jemth, P. (2021). CRISPR/Cas9-based inactivation of human papillomavirus oncogenes E6 or E7 induces senescence in cervical cancer cells. Virology 562, 92–102. 10.1016/j.virol.2021.07.003.

46. Janiszewska, J., Kostrzewska-Poczekaj, M., Wierzbicka, M., Brenner, J.C., and Giefing, M. (2025). HPV-driven oncogenesis: much more than the E6 and E7 oncoproteins. J. Appl. Genet. 66, 63–71. 10.1007/s13353-024-00861-2.

47. Letafati, A., Shahbazi, S., Zarei, M., and Ghasemi, F. (2024). Emerging paradigms: unmasking the role of oxidative stress in HPV-induced carcinogenesis. Infect. Agent. Cancer 19, 30. 10.1186/s13027-024-00574-9.

48. Münger, K., Phelps, W.C., Bubb, V., Howley, P.M., and Schlegel, R. (1989). The E6 and E7 genes of the human papillomavirus type 16 together are necessary and sufficient for transformation of primary human keratinocytes. J. Virol. 63, 4417–4421. 10.1128/JVI.63.10.4417-4421.1989.

49. Ranasinghe, V., and McMillan, N.A.J. (2025). Novel therapeutic strategies for targeting E6 and E7 oncoproteins in cervical cancer. Crit. Rev. Oncol. Hematol. 211, 104721. 10.1016/j.critrevonc.2025.104721.

50. Fert, I., Nguyen, J., Decoster, G., Nguyen, T., and Goubier, A. (2024). T-cell immunity induced and reshaped by an anti-HPV immuno-oncotherapeutic lentiviral vector. NPJ Vaccines 9, 102. 10.1038/s41541-024-00902-6.

51. Lee, S., Kim, J., Park, H., Choi, Y., and Kim, Y. (2023). mRNA-HPV vaccine encoding E6 and E7 improves therapeutic potential for HPV-mediated cancers via subcutaneous immunization. J. Med. Virol. 95, e29309. 10.1002/jmv.29309.

52. Zhang, Y., Qiu, K., Ai, J., Xu, M., Wang, B., Alu, A., et al. (2025). Ad-E6/7-HR vaccine improves the prophylactic and therapeutic efficacy in HPV-associated cancers. Clin. Transl. Med. 15, e70305. 10.1002/ctm2.70305.

53. Tsukamoto, K., Yamashita, A., Maeki, M., Tokeshi, M., Imai, H., Fukao, A., et al. (2024). Enhanced broad-spectrum efficacy of an L2-based mRNA vaccine targeting HPV types 6, 11, 16, 18, with cross-protection against multiple additional high-risk types. Vaccines 12, 1239. 10.3390/vaccines12111239.

54. Wang, J.W., and Roden, R.B.S. (2013). L2, the minor capsid protein of papillomavirus. Virology 445, 175–186. 10.1016/j.virol.2013.04.017.

55. Zhao, X., Yang, F., Mariz, F., Osen, W., Bolchi, A., Ottonello, S., et al. (2020). Combined prophylactic and therapeutic immune responses against human papillomaviruses induced by a thioredoxin-based L2-E7 nanoparticle vaccine. PLOS Pathog. 16, e1008827. 10.1371/journal.ppat.1008827.

56. Sadeghi, Z., Aboofazeli, A., Sarrafzadeh, S., Tayebi, N., Tajdini, R., Yarandi, F., et al. (2025). Investigating the link between HPV genotypes and cervical abnormality incidence in women with HPV infections: insights from a leading referral centre. Virol. J. 22, 264. 10.1186/s12985-025-02858-9.

57. Jemon, K., Leong, C.M., Ly, K., Young, S.L., McLellan, A.D., and Hibma, M.H. (2016). Suppression of the CD8 T cell response by human papillomavirus type 16 E7 occurs in Langerhans cell-depleted mice. Sci. Rep. 6, 34789. 10.1038/srep34789.

58. Maskey, N., Thapa, N., Maharjan, M., Shrestha, G., Maharjan, N., Cai, H., et al. (2019). Infiltrating CD4 and CD8 lymphocytes in HPV infected uterine cervical milieu. Cancer Manag. Res. 11, 7647–7655. 10.2147/CMAR.S217264.

59. Mondatore, D., Bai, F., Augello, M., Giovenzana, M., Pisani Ceretti, A., Bono, V., et al. (2022). Persistence of high percentage of peripheral activated CD8+ T cells predict cytologic HPV-related dysplasia in cART-treated, HIV-positive subjects. Open Forum Infect. Dis. 9, ofac046. 10.1093/ofid/ofac046.

60. Bhattacharjee, R., Das, S.S., Biswal, S.S., Nath, A., Das, D., Basu, A., et al. (2022). Mechanistic role of HPV-associated early proteins in cervical cancer: molecular pathways and targeted therapeutic strategies. Crit. Rev. Oncol. Hematol. 174, 103675. 10.1016/j.critrevonc.2022.103675.

61. Wang, C.Y., Peng, W.J., Kuo, B.S., Ho, Y.H., Wang, M.S., Yang, Y.T., et al. (2023). Toward a pan-SARS-CoV-2 vaccine targeting conserved epitopes on spike and non-spike proteins for potent, broad and durable immune responses. PLOS Pathog. 19, e1010870. 10.1371/journal.ppat.1010870.

62. Medhasi, S., and Chantratita, N. (2022). Human leukocyte antigen (HLA) system: genetics and association with bacterial and viral infections. J. Immunol. Res. 2022, 9710376. 10.1155/2022/9710376.

63. Ahmels, M., Mariz, F.C., Braspenning-Wesch, I., Stephan, S., Huber, B., Schmidt, G., et al. (2022). Next generation L2-based HPV vaccines cross-protect against cutaneous papillomavirus infection and tumor development. Front. Immunol. 13, 1010790. 10.3389/fimmu.2022.1010790.

64. Hancock, G., Blight, J., Lopez-Camacho, C., Kopycinski, J., Pocock, M., Byrne, W., et al. (2019). A multi-genotype therapeutic human papillomavirus vaccine elicits potent T cell responses to conserved regions of early proteins. Sci. Rep. 9, 18713. 10.1038/s41598-019-55014-z.

65. Riemer, A.B., Keskin, D.B., Zhang, G., Handley, M., Anderson, K.S., Brusic, V., et al. (2010). A conserved E7-derived cytotoxic T lymphocyte epitope expressed on human papillomavirus 16-transformed HLA-A2+ epithelial cancers. J. Biol. Chem. 285, 29608–29622. 10.1074/jbc.M110.126722.

66. Grabowska, A.K., Kaufmann, A.M., and Riemer, A.B. (2015). Identification of promiscuous HPV16-derived T helper cell epitopes for therapeutic HPV vaccine design. Int. J. Cancer 136, 212–224. 10.1002/ijc.28968.

67. Rock, K.L., Reits, E., and Neefjes, J. (2016). Present yourself! By MHC class I and MHC class II molecules. Trends Immunol. 37, 724–737. 10.1016/j.it.2016.08.010.

